# Blood lead levels and bladder cancer among US participants: NHANES 1999–2018

**DOI:** 10.1101/2024.02.22.24303198

**Authors:** Mei Huang, Hongxiao Li, Jiahui Chen, Liuqiang Li, Yifei Zhan, Jun Bian, Meiling Chen, Dehui Lai

**Author notes:** These authors contributed equally to this work (Mei Huang, Hongxiao Li). **Correspondent:** Dehui Lai Jun Bian Meiling Chen, 621 Gangwan Road, Huangpu District, Guangzhou City, Guangdong Province. **Conflict of Interest** The authors declare they have no actual or potential competing financial interests.

## Abstract

**Background:** Lead is a toxic metal for human health, but its carcinogenicity is controversial, and the effect on bladder cancer is still unknown. The object of this study was to demonstrate the link between blood lead and bladder cancer.

**Objectives:** We investigated associations of lead exposures with bladder cancer.

**Methods:** We used the database from the National Health and Nutrition Examination Survey (NHANES, 1999–2018) to perform a cross-sectional study. We performed Weighted multivariate logistic regression to examine the association between blood lead level with bladder cancer, and then a subgroup analysis was performed. The nonlinear association between BLL and bladder cancer was described using fitted smoothing curves.

**Results:** A total of 40,486 participants were included in this study, the mean (SD) BMI was 28.71 ± 6.68 kg/m^2^. A fully adjusted model showed that BLL was parallel associated with bladder cancer (Odds ratio [OR] = 2.946, 95% Confidence interval [CI] = 1.025 to 8.465, P = 0.047) in people with BMI < 28kg/m^2^. However, no difference was found in BMI≥28kg/m2 subgroup and in General population. In subgroup analysis of participants with BMI < 28kg/m^2^, blood lead was associated with bladder cancer in the male, non-hypertensive, < 70 year old subgroup (*p* < 0.05), but was not significantly different from the other subgroups. Additionally, we discovered a non-linear association between BLL and bladder cancer using a linear regression model.

**Discussion:** In this cross-sectional study, we identified higher BLL level was independently associated with Bladder cancer in People with BMI<28kg/m^2^.The results compensated for earlier investigations, but more large-scale prospective cohorts were required for validation.

## Introduction

Lead is a heavy metal that can cause environmental pollution and is poisonous. It is widely found in life, such as mining, welding, batteries, making stained glass, ceramics, especially in low - and middle-income countries(Mitra et al. 2017; Obeng-Gyasi 2019; Walter 2023). Lead is difficult to decompose and can be divided into organic and inorganic forms. It mainly enters the human body through digestion, respiration and skin(Fu and Xi 2020). The absorption rate can vary from 10-50% according to the state of the exposed person and the physicochemical factors of lead(Huang 2022). 5mg/dl for adults and 3.5ug/dl for children was defined as reference value, but the threshold of harmful concentration is not clear(Ruckart et al. 2021). Lead affects cell metabolism by affecting antioxidant reaction, key enzymes and various hormone activities. Oxidative stress reactions caused by heavy metals can also cause cancer by interfering with DNA repair (Dhir et al. 2011; Flora et al. 2008; Genestra 2007). Recently, with the progress of social industrialization, human health is increasingly affected by heavy metals(Rusyniak et al. 2010).

Bladder cancer is the second most common malignancy of the urinary system and ranks 10th in absolute incidence worldwide, with 549,000 new cases and approximately 200,000 deaths annually. In the United States, bladder cancer is the sixth highest incidence of cancer. Urothelial carcinoma is the main type of bladder cancer, accounting for more than 90%. Of all cancers, bladder cancer has the highest lifetime treatment costs, with total annual treatment costs of around €3.6 billion in the United States and nearly €5 billion in Europe, adding to the burden on the global economy. Smoking is the strongest risk factor, but occupational and environmental toxins also significantly increase the disease burden of bladder cancer(Compérat et al. 2022; Richters et al. 2020; Saginala et al. 2020).

There are many reports about the harm of lead to human body, such as type 2 diabetes(Zhu et al. 2022), urinary incontinence(Ni et al. 2022), hypertension(Huang 2022; Tang et al. 2022), cirrhosis of liver(Reja et al. 2020), lung cancer(Rhee et al. 2021), kidney function, Neuropsychiatric problems and endocrine diseases(Qayyum et al. 2012; Walter 2023), but there are few reports about the correlation between lead and bladder cancer, whether lead is a risk factor for bladder cancer is still controversial. This study aims to reveal the relationship between the lead and bladder cancer.

## Methods

### Study population

NHANES is a nationally representative survey which was designed to assess the health and nutrition status of adults and children in the United States. Socioeconomic status, diet, medical and physiological examinations and laboratory tests, were conducted on the participants. The database for this article is available from this website (https://www.cdc.gov/nchs/NHANES/index.htm). We analyzed the data from the NHANES survey cycles (1999–2018). In short, To explore a nationally representative sample, NHANES adopts a complicated, multistage probability sampling approach among the civilian, noninstitutionalized US population. All participants completed household questionnaires administered by trained research personnel, which included demographic and health history questions. Standardized medical examinations, blood sample collection, and other in-person testing at mobile examination facilities are also part of the research procedure. All NHANES participants provided informed consent. During 1999–2018 NHANES cycles, 101,316 participants were investigated. There were 25,555 participants without blood lead data. Participants without cancer data or had other cancers were excluded in this study(n=35275). The final study population of this study was 40,486. The process of recruiting is shown in Figure 1.

**Figure 1.**
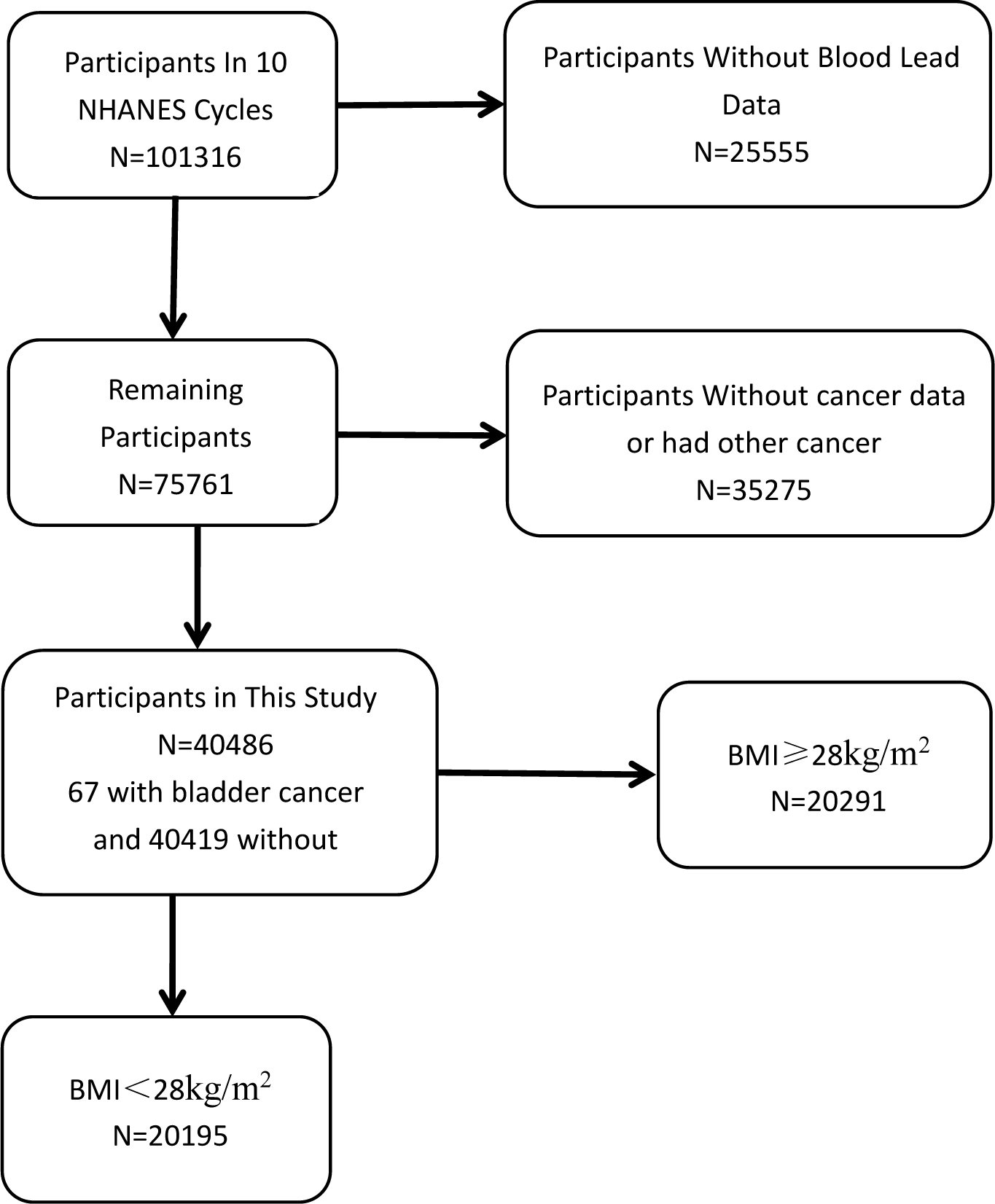
The flowchat of participants.

### Study Variable

#### Blood Lead

The blood lead levels were examined at the Environmental Health Sciences Laboratory of the Center for Disease Control and Prevention (CDC) National Center for Environmental Health (NCEH). To assess the blood lead level, venous blood was obtained. Before measuring, blood samples were simply diluted and kept at −20°C. The blood lead level was measured in the central laboSpectrometer (ELAN DRC II, PerkinElmer, Norwalk) in accordance with the usual meratory using an Inductively Coupled Plasma Dynamic Reaction Cell Mass thodology. The lower limits of detection (LOD) for blood lead were different in each NHANES cycle, which are summarized in Table 1. The detection rates of blood lead for participants is 99.7%. All blood lead levels less than the lower limit of detection were replaced with the lower limit of detection multiplied by √2. The specific laboratory method, detection limit, and other data can be obtained on the official website.

**Table 1.** The limit of detection (LOD) for lead in each NHANES cycle.

| NHANES cycle | lead, ug/dL |
| --- | --- |
| 2017-2018 | 0.07 |
| 2015-2016 | 0.07 |
| 2013-2014 | 0.07 |
| 2011-2012 | 0.25 |
| 2009-2010 | 0.18 |
| 2007-2008 | 0.26 |
| 2005-2006 | 0.25 |
| 2003-2004 | 0.25 |
| 2001-2002 | 0.20 |
| 1999-2000 | 0.20 |

#### Bladder cancer

The answer to the question “Ever been told you had cancer or malignancy?” on the MCQ questionnaire was used for learning whether the participants had cancer or malignancy. The answer to the question “What kind of cancer was it?” gave the location of cancer.

#### Other clinical Variable

Data on age, sex, race, education, marital, ratio of family income to poverty (PIR), BMI, alcohol, high blood pressure, diabetes, creatinine level, smoke were also collected. Age, sex, race, education, marriage, and PIR can be obtained from demographic information (DEMO). The information of hypertension, diabetes, smoking and drinking can be obtained from questionnaires BPQ, DIQ, SMQ and QLQ respectively. BMI was obtained in the physical examination and blood creatinine in the column of laboratory examination. More details of these covariate data can be found in www.cdc.gov/nchs/NHANES/. Mean values are used when the covariates of continuous variables are missing(PIR 2.98, BMI 28.71, blood creatine 77.10 umol/L). The classification of all variables is shown in Table 2.

**TABLE 2.**
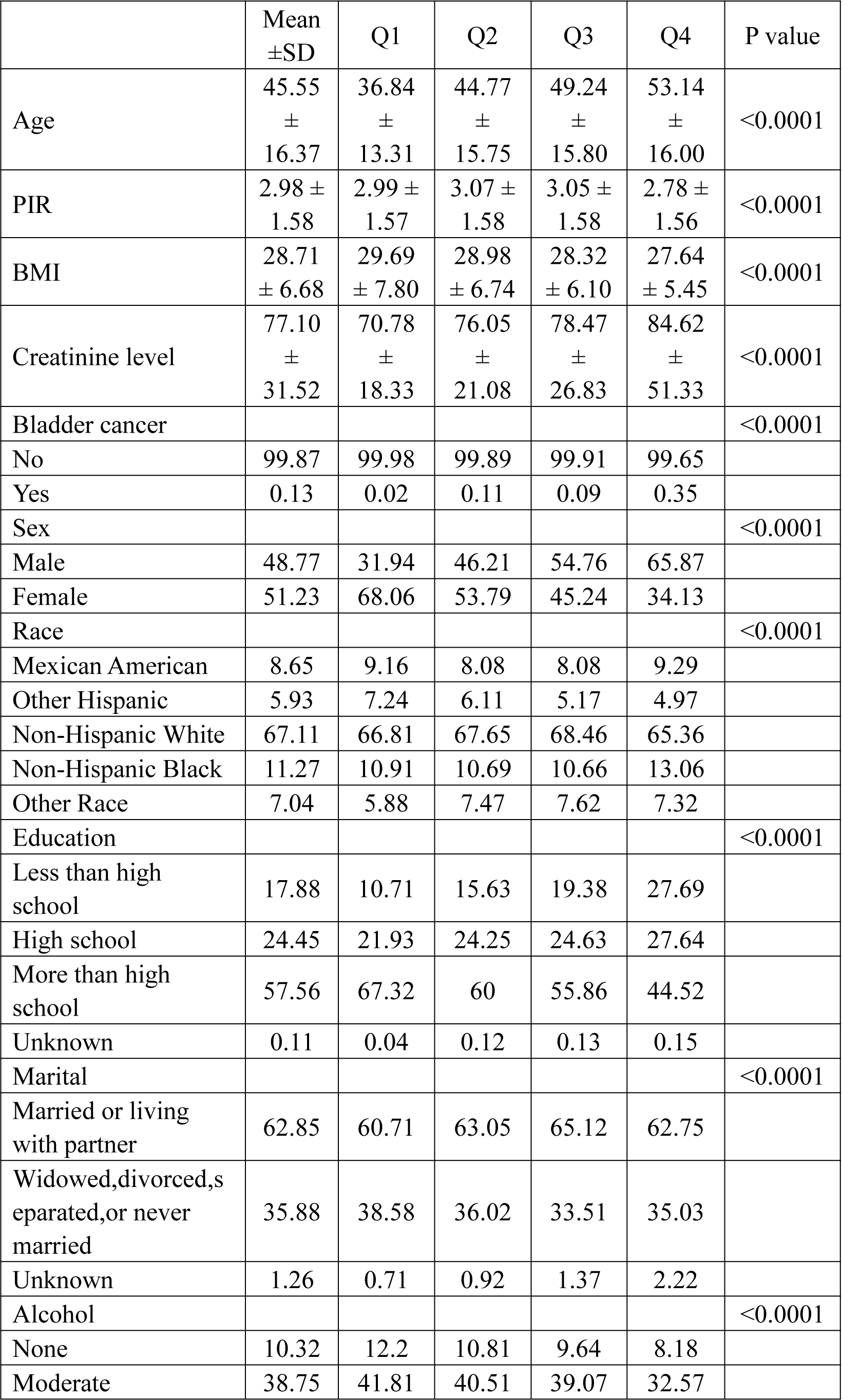
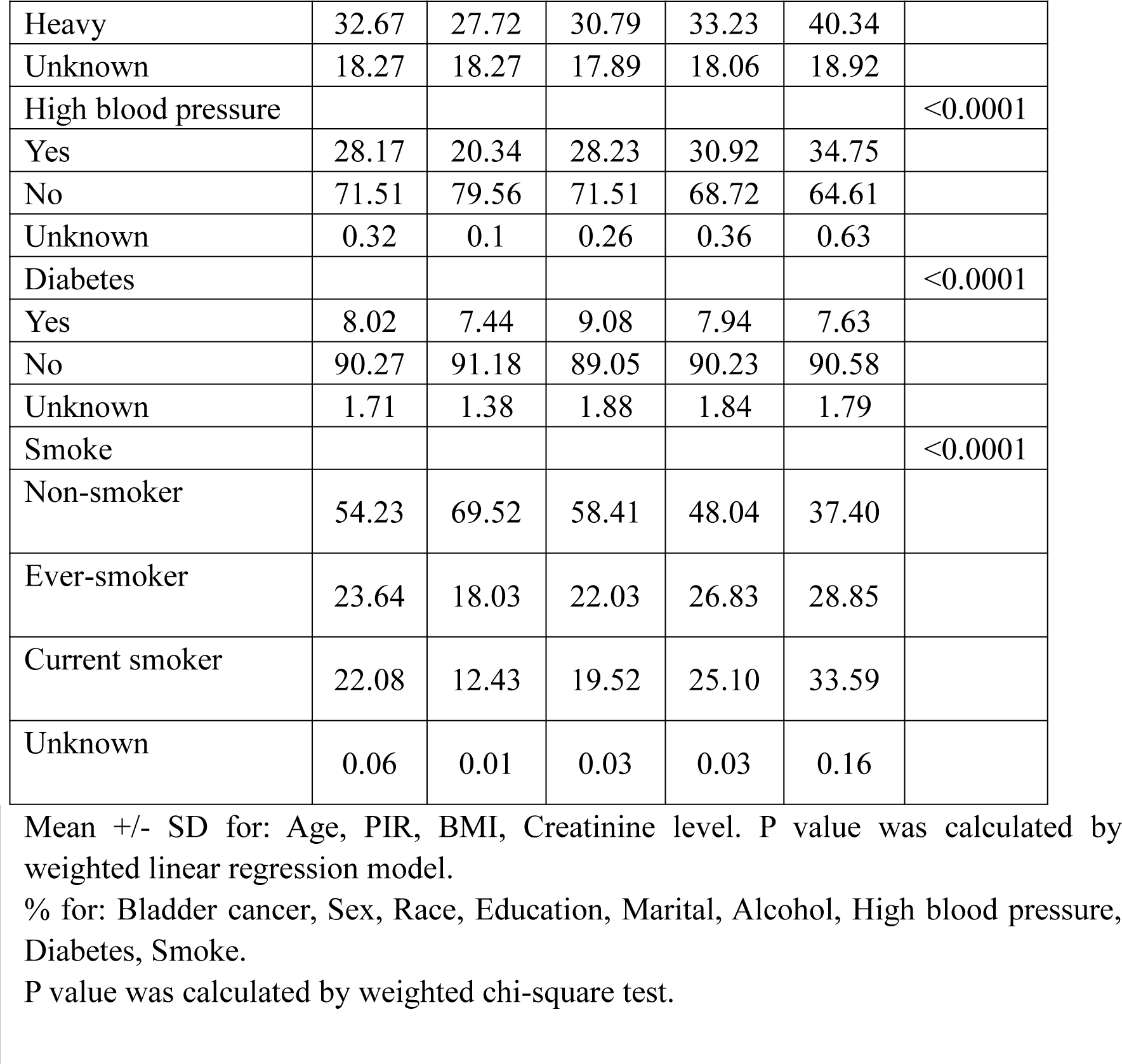
| Weighted characteristics of the study population based on serum lead.

|  | Mean<br>±SD | Q1 | Q2 | Q3 | Q4 | P value |
| --- | --- | --- | --- | --- | --- | --- |
| Age | 45.55<br>±<br>16.37 | 36.84<br>±<br>13.31 | 44.77<br>±<br>15.75 | 49.24<br>±<br>15.80 | 53.14<br>±<br>16.00 | <0.0001 |
| PIR | 2.98 ±<br>1.58 | 2.99 ±<br>1.57 | 3.07 ±<br>1.58 | 3.05 ±<br>1.58 | 2.78 ±<br>1.56 | <0.0001 |
| BMI | 28.71<br>± 6.68 | 29.69<br>± 7.80 | 28.98<br>± 6.74 | 28.32<br>± 6.10 | 27.64<br>± 5.45 | <0.0001 |
| Creatinine level | 77.10<br>±<br>31.52 | 70.78<br>±<br>18.33 | 76.05<br>±<br>21.08 | 78.47<br>±<br>26.83 | 84.62<br>±<br>51.33 | <0.0001 |
| Bladder cancer |  |  |  |  |  | <0.0001 |
| No | 99.87 | 99.98 | 99.89 | 99.91 | 99.65 |  |
| Yes | 0.13 | 0.02 | 0.11 | 0.09 | 0.35 |  |
| Sex |  |  |  |  |  | <0.0001 |
| Male | 48.77 | 31.94 | 46.21 | 54.76 | 65.87 |  |
| Female | 51.23 | 68.06 | 53.79 | 45.24 | 34.13 |  |
| Race |  |  |  |  |  | <0.0001 |
| Mexican American | 8.65 | 9.16 | 8.08 | 8.08 | 9.29 |  |
| Other Hispanic | 5.93 | 7.24 | 6.11 | 5.17 | 4.97 |  |
| Non-Hispanic White | 67.11 | 66.81 | 67.65 | 68.46 | 65.36 |  |
| Non-Hispanic Black | 11.27 | 10.91 | 10.69 | 10.66 | 13.06 |  |
| Other Race | 7.04 | 5.88 | 7.47 | 7.62 | 7.32 |  |
| Education |  |  |  |  |  | <0.0001 |
| Less than high school | 17.88 | 10.71 | 15.63 | 19.38 | 27.69 |  |
| High school | 24.45 | 21.93 | 24.25 | 24.63 | 27.64 |  |
| More than high school | 57.56 | 67.32 | 60 | 55.86 | 44.52 |  |
| Unknown | 0.11 | 0.04 | 0.12 | 0.13 | 0.15 |  |
| Marital |  |  |  |  |  | <0.0001 |
| Married or living with partner | 62.85 | 60.71 | 63.05 | 65.12 | 62.75 |  |
| Widowed,divorced,s eparated,or never married | 35.88 | 38.58 | 36.02 | 33.51 | 35.03 |  |
| Unknown | 1.26 | 0.71 | 0.92 | 1.37 | 2.22 |  |
| Alcohol |  |  |  |  |  | <0.0001 |
| None | 10.32 | 12.2 | 10.81 | 9.64 | 8.18 |  |
| Moderate | 38.75 | 41.81 | 40.51 | 39.07 | 32.57 |  |
| Heavy | 32.67 | 27.72 | 30.79 | 33.23 | 40.34 |  |
| Unknown | 18.27 | 18.27 | 17.89 | 18.06 | 18.92 |  |
| High blood pressure |  |  |  |  |  | <0.0001 |
| Yes | 28.17 | 20.34 | 28.23 | 30.92 | 34.75 |  |
| No | 71.51 | 79.56 | 71.51 | 68.72 | 64.61 |  |
| Unknown | 0.32 | 0.1 | 0.26 | 0.36 | 0.63 |  |
| Diabetes |  |  |  |  |  | <0.0001 |
| Yes | 8.02 | 7.44 | 9.08 | 7.94 | 7.63 |  |
| No | 90.27 | 91.18 | 89.05 | 90.23 | 90.58 |  |
| Unknown | 1.71 | 1.38 | 1.88 | 1.84 | 1.79 |  |
| Smoke |  |  |  |  |  | <0.0001 |
| Non-smoker | 54.23 | 69.52 | 58.41 | 48.04 | 37.40 |  |
| Ever-smoker | 23.64 | 18.03 | 22.03 | 26.83 | 28.85 |  |
| Current smoker | 22.08 | 12.43 | 19.52 | 25.10 | 33.59 |  |
| Unknown | 0.06 | 0.01 | 0.03 | 0.03 | 0.16 |  |

Mean +/- SD for: Age, PIR, BMI, Creatinine level. P value was calculated by weighted linear regression model.
% for: Bladder cancer, Sex, Race, Education, Marital, Alcohol, High blood pressure, Diabetes, Smoke.
P value was calculated by weighted chi-square test.

#### Statistical Analyses

All the analyses were performed with R (version 3.4.3, http://www.R-project.org) and Empower Stats software (http://www.empowerstats.com), Stata/MP16.0. Continuous variables were reported as mean ± standard deviation (SD), while the categorical variables were percentages. Baseline characteristics were analyzed by a linear regression model for continuous variables and a chi-square test for categorical variables, respectively. We divided the blood lead concentration into four categories. Weighted multivariate logistic regression models were performed to explore the independent association of blood lead concentration with bladder cancer after adjusting for potential confounding factors. Three models were built: Model 1, an unadjusted model; Model 2, minimally adjusted model (adjusted for age, sex, race, education, marital and PIR), and Model 3, a fully adjusted model (adjusted for age, sex, race, education, marital and PIR, BMI, alcohol use, high blood pressure, diabetes and creatinine level, smoke). BMI-stratified subgroup analysis was performed and Further stratified logistic regression analysis was conducted to clarify the association between Blood lead level (BLL) and bladder cancer. Multivariate tests were constructed by controlling for variables and fitting a smooth curve. Subgroup analyses were performed and they were stratified by sex, age, high blood pressure and PIR. The value of p < 0.05 was considered statistically significant.

## Results

### Baseline Characteristic

The clinical characteristics of the participants according to blood lead level as a column stratified variable are shown in Table 2. A total of 40,486 participants aged 20–85 years who had measured data for blood lead and bladder cancer were included in this study. Of all the participants, there were 48.77% were men, and the mean age was 45.55 ± 16.37. Participants had an average BMI of 28.71, the gender ratio was roughly equal, and more than half of the participants were non-Hispanic white, had a high school education or more, and were married or living with a partner. Most people did not have high blood pressure and diabetes, but most people had a history of alcohol consumption, mostly moderate alcohol consumption. More than half of the participants were non-smokers. There were 0.17% of the participants with bladder cancer.

Participants who fell into the Quartile 4 group tended to be older, male, lower PIR and BMI, higher creatinine level, Non-Hispanic white, married or living with a partner, heavy alcohol consumption, smoker, without hypertension or diabetes. Besides, participants who are older, male, lower PIR and BMI, higher creatinine level, high school diploma or below, heavy alcohol consumption, former or current smoker, with hypertension and bladder cancer patients are more likely to have higher blood lead level.

We can draw the following conclusions from the Table 2: Blood lead levels were associated with age, sex, education, PIR, BMI, creatinine levels, smoking and alcohol consumption, bladder cancer and high blood pressure (p<0.001). The correlation between blood lead and blood pressure is consistent with the results reported by predecessors

### The Association of Blood Lead Concentration With bladder cancer

Table 3 showed the results of the multivariable regression analysis between BLL and bladder cancer. In unadjusted model 1, we observed statistically significant associations between elevated BLLs and bladder cancer (OR 6.350, 95% CI 4.048, 9.961, P<0.001). The risk of bladder cancer in the highest quartile array was 16.387 times higher than in the lowest group. After adjusting covariates, however, these associations weakened substantially and were no longer statistically significant in model 2 (OR 3.316, 95% CI 1.161, 9.437, P= 0.027) and model3 (OR 2.280,95% CI 0.575,9.038, P=0.243). When adjusting BMI<28kg/m^2^ (Table 4), the univariate and multivariate analyses demonstrated high BLL was associated with a higher risk of bladder cancer in the crude model (OR 7.379, 95% CI 3.959,13.751, P < 0.001), model 2 (OR 3.649, 95% CI 1.380,9.649, P=0.010), and modle 3(OR 2.946, 95% CI 1.025,8.465, P=0.047). We can draw a conclusion from Table 4 that after adjusting for all confounding factors, BLLs was still positively associated with bladder cancer in the population with BMI< 28kg/m^2^ (P=0.047). The correlation was not found when weighted multivariate logistic regression analysis was conducted in BMI≥28kg/m^2^ group, suggesting that no potential modifiers in the relationship between BLLs and bladder cancer in obese population (Table 5).

**TABLE 3.** | Association between blood lead levels (μg/dl) and bladder cancer.

|  | Model 1 | Model 2 | Model 3 |
| --- | --- | --- | --- |
|  | OR(95% CI) P<br>value | OR(95% CI) P<br>value | OR(95% CI) P<br>value |
| Serum<br>Lead(umol/L) | 6.350<br>(4.048,9.961)<0.001 | 3.316<br>(1.161,9.473)<br>0.027 | 2.280<br>(0.575,9.038)0.243 |
| Serum Lead<br>categories |  |  |  |
| Q1 (0.000-0.039<br>umol/L) | Reference | Reference | Reference |
| Q2 (0.040-0.062<br>umol/L) | 4.959<br>(1.072,22.943)<br>0.042 | 1.821<br>(0.376,8.833)<br>0.458 | 1.861 (0.392,8.841)<br>0.436 |
| Q3 (0.063-0.100<br>umol/L) | 4.076<br>(0.936,17.737)<br>0.063 | 0.912<br>(0.205,4.059)<br>0.904 | 0.767 (0.171,3.438)<br>0.729 |
| Q4 (0.101-2.960<br>umol/L) | 16.387<br>(3.681,72.957)<<br>0.001 | 2.260<br>(0.460,11.095)<br>0.317 | 1.730 (0.367,8.146)<br>0.490 |
| P for trend | <0.001 | 0.234 | 0.379 |

Model 1: No covariates were adjusted.
Model 2:Age, Sex, Race, Education, Marital, PIR
Model 3:Age, Sex, Race, Education, Marital, PIR, BMI, Alcohol, High blood pressure,
Diabetes, Creatinine level, Smoke.

**TABLE 4.**
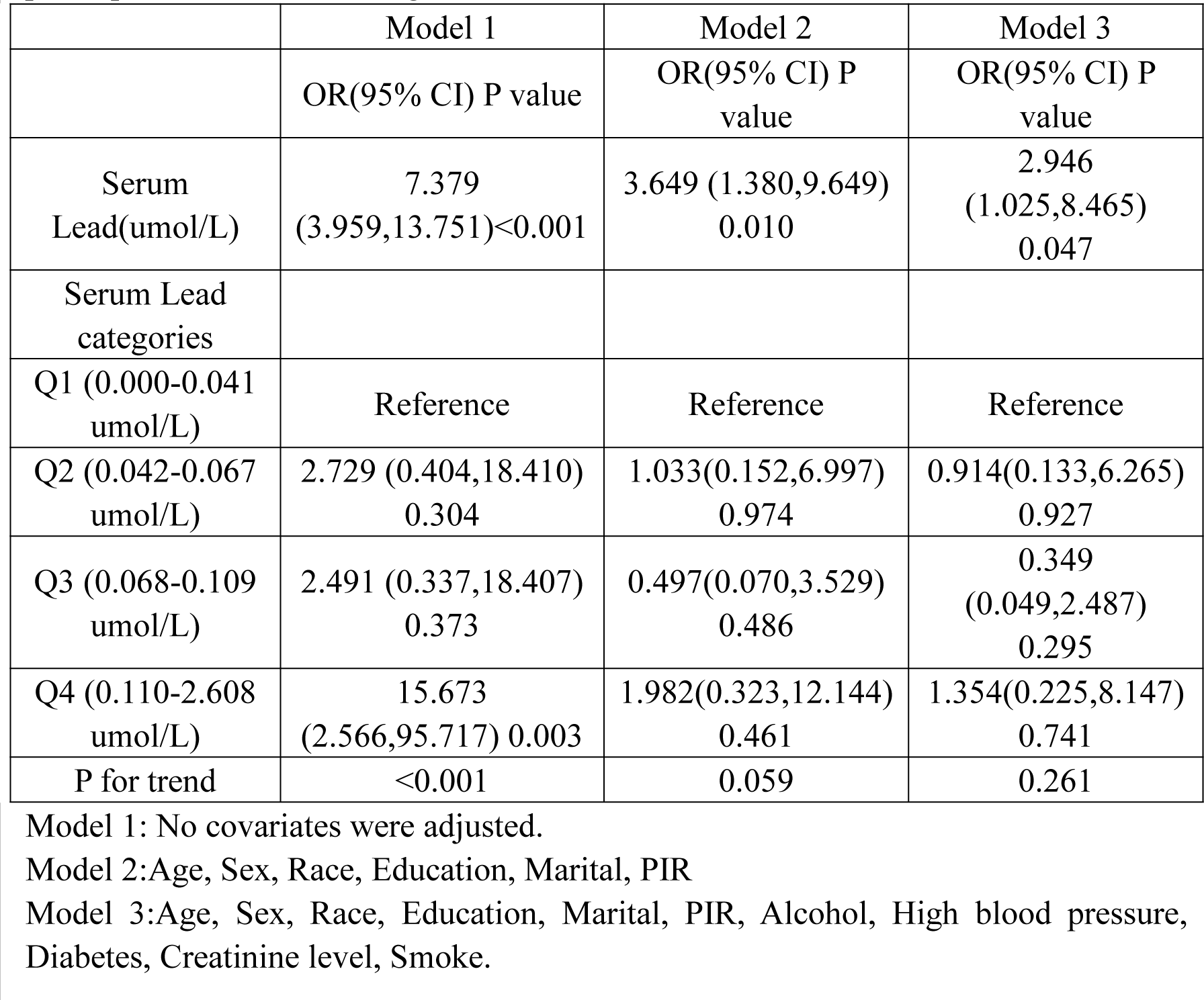
| Association between blood lead levels (μg/dl) and bladder cancer in participants with BMI<28kg/m^2^.

|  | Model 1 | Model 2 | Model 3 |
| --- | --- | --- | --- |
|  | OR(95% CI) P value | OR(95% CI) P value | OR(95% CI) P value |
| Serum Lead($\mu\text{mol/L}$) | 7.379<br>(3.959,13.751)<0.001 | 3.649 (1.380,9.649)<br>0.010 | 2.946<br>(1.025,8.465)<br>0.047 |
| Serum Lead categories |  |  |  |
| Q1 (0.000-0.041 $\mu\text{mol/L}$) | Reference | Reference | Reference |
| Q2 (0.042-0.067 $\mu\text{mol/L}$) | 2.729 (0.404,18.410)<br>0.304 | 1.033(0.152,6.997)<br>0.974 | 0.914(0.133,6.265)<br>0.927 |
| Q3 (0.068-0.109 $\mu\text{mol/L}$) | 2.491 (0.337,18.407)<br>0.373 | 0.497(0.070,3.529)<br>0.486 | 0.349<br>(0.049,2.487)<br>0.295 |
| Q4 (0.110-2.608 $\mu\text{mol/L}$) | 15.673<br>(2.566,95.717) 0.003 | 1.982(0.323,12.144)<br>0.461 | 1.354(0.225,8.147)<br>0.741 |
| P for trend | <0.001 | 0.059 | 0.261 |

Model 1: No covariates were adjusted.
Model 2: Age, Sex, Race, Education, Marital, PIR
Model 3: Age, Sex, Race, Education, Marital, PIR, Alcohol, High blood pressure, Diabetes, Creatinine level, Smoke.

**TABLE 5.** | Association between blood lead levels (μg/dl) and bladder cancer in participants with BMI≥28kg/m^2^.

|  | Model 1 | Model 2 | Model 3 |
| --- | --- | --- | --- |
|  | OR(95% CI) P value | OR(95% CI) P value | OR(95% CI) P value |
| Serum Lead($\mu\text{mol/L}$) | 4.999<br>(2.677,9.336)<0.001 | 1.026<br>(0.023,45.896)<br>0.990 | 0.303<br>(0.003,33.968)<br>0.621 |
| Serum Lead categories |  |  |  |
| Q1<br>(0.0000-0.0380) | Reference | Reference | Reference |
| Q2<br>(0.0390-0.0590) | 5.771(0.511,65.194)<br>0.159 | 2.578(0.189,35.186)<br>0.479 | 2.350(0.189,29.188)<br>0.507 |
| Q3<br>(0.0600-0.0960) | 12.195<br>(1.473,100.942)<br>0.022 | 3.406(0.368,31.551)<br>0.282 | 2.679<br>(0.297,24.202)<br>0.382 |
| Q4<br>(0.0970-2.9600) | 21.785<br>(2.655,178.747)<br>0.005 | 3.944(0.378,41.179)<br>0.254 | 2.577(0.294,22.563)<br>0.394 |
| P for trend | <0.001 | 0.3126 | 0.3102 |

Model 1: No covariates were adjusted.
Model 2: Age, Sex, Race, Education, Marital, PIR
Model 3: Age, Sex, Race, Education, Marital, PIR, Alcohol, High blood pressure, Diabetes, Creatinine level, Smoke.

The weighted characteristics of the population with BMI<28kg/m^2^ are shown in Table 6.

**TABLE 6.** | Weighted characteristics of the study population based on blood lead in participants with BMI<28kg/m^2^.

|  | Mean<br>±SD | Q1 | Q2 | Q3 | Q4 | P value |
| --- | --- | --- | --- | --- | --- | --- |
| Age | 44.09<br>±<br>16.85 | 35.14<br>±<br>13.20 | 42.73<br>±<br>16.14 | 48.56<br>±<br>16.30 | 52.54<br>±<br>16.68 | <0.0001 |
| PIR | 3.03 ±<br>1.59 | 3.11 ±<br>1.58 | 3.16 ±<br>1.60 | 3.08 ±<br>1.59 | 2.70 ±<br>1.54 | <0.0001 |
| Creatinine level | 76.30<br>±<br>31.85 | 70.95<br>±<br>17.13 | 75.06<br>±<br>24.52 | 77.32<br>±<br>23.37 | 83.96<br>±<br>54.95 | <0.0001 |
| Bladder cancer |  |  |  |  |  | <0.0001 |
| No | 99.86 | 99.97 | 99.92 | 99.93 | 99.56 |  |
| Yes | 0.14 | 0.03 | 0.08 | 0.07 | 0.44 |  |
| Sex |  |  |  |  |  | <0.0001 |
| Male | 48.31 | 31.97 | 44.89 | 54.81 | 67.02 |  |
| Female | 51.69 | 68.03 | 55.11 | 45.19 | 32.98 |  |
| Race |  |  |  |  |  | <0.0001 |
| Mexican American | 7.54 | 7.57 | 6.88 | 7.05 | 8.94 |  |
| Other Hispanic | 5.72 | 7.06 | 5.60 | 4.97 | 4.97 |  |
| Non-Hispanic White | 68.72 | 69.28 | 70.75 | 69.68 | 64.16 |  |
| Non-Hispanic Black | 9.13 | 8.68 | 7.74 | 8.36 | 12.49 |  |
| Other Race | 8.88 | 7.40 | 9.02 | 9.94 | 9.43 |  |
| Education |  |  |  |  |  | <0.0001 |
| Less than high school | 17.09 | 8.63 | 14.80 | 19.01 | 29.26 |  |
| High school | 22.52 | 19.50 | 22.31 | 22.28 | 27.24 |  |
| More than high school | 60.28 | 71.77 | 62.84 | 58.60 | 43.32 |  |
| Unknown | 0.11 | 0.10 | 0.05 | 0.11 | 0.18 |  |
| Marital |  |  |  |  |  | <0.0001 |
| Married or living with partner | 61.59 | 58.63 | 62.44 | 64.46 | 60.98 |  |
| Widowed,divorced,separated,or never married | 37.03 | 40.71 | 36.42 | 33.97 | 36.56 |  |
| Unknown | 1.39 | 0.67 | 1.14 | 1.57 | 2.46 |  |
| Alcohol |  |  |  |  |  | <0.0001 |
| None | 10.10 | 11.95 | 10.89 | 9.04 | 7.89 |  |
| Moderate | 41.29 | 45.23 | 42.71 | 41.62 | 33.64 |  |
| Heavy | 32.15 | 26.35 | 30.03 | 33.14 | 41.61 |  |
| Unknown | 16.45 | 16.46 | 16.37 | 16.21 | 16.86 |  |
| High blood pressure |  |  |  |  |  | <0.0001 |
| Yes | 19.05 | 10.56 | 17.93 | 22.67 | 27.66 |  |
| No | 80.57 | 89.39 | 81.83 | 76.85 | 71.48 |  |
| Unknown | 0.38 | 0.05 | 0.24 | 0.49 | 0.86 |  |
| Diabetes |  |  |  |  |  | 0.0010 |
| Yes | 4.31 | 3.61 | 4.16 | 4.63 | 5.04 |  |
| No | 94.70 | 95.70 | 94.78 | 94.28 | 93.74 |  |
| Unknown | 0.99 | 0.68 | 1.05 | 1.09 | 1.22 |  |
| Smoke |  |  |  |  |  | <0.0001 |
| Non-smoker | 53.86 | 71.73 | 57.71 | 45.19 | 35.18 |  |
| Non-smoker | 21.29 | 15.63 | 20.81 | 25.24 | 24.74 |  |
| Current smoker | 24.78 | 12.63 | 21.42 | 29.54 | 39.85 |  |
| Unknown | 0.07 | 0.01 | 0.05 | 0.03 | 0.23 |  |

Mean +/- SD for: Age, PIR, Creatinine level. P value was calculated by weighted linear regression model.
% for: Bladder cancer, Sex, Race, Education, Marital, Alcohol, High blood pressure, Diabetes, Smoke. P value was calculated by weighted chi-square test.

Further subgroup analysis of participants with BMI < 28kg/m^2^ was conducted, the result is shown in Table 7. Blood lead was associated with bladder cancer in the male (P=0.01), less than 70 year old (P<0.01), non-hypertensive (P=0.01) subgroup, but was not significantly different from the other subgroups.

**Table 7.** | Subgroup analyses of the association between blood lead and bladder cancer in participants with BMI<28kg/m^2^.

|  | OR | 95%CI<br>low | 95%CI upp | P.value |
| --- | --- | --- | --- | --- |
| Sex |  |  |  |  |
| Male | 3.64 | 1.32 | 10.06 | 0.01 |
| Female | 0.00 | 0.00 | 15895.88 | 0.43 |
| Age |  |  |  |  |
| <70 years | 6.50 | 2.12 | 19.93 | 0.00 |
| >=70 years | 3.21 | 0.80 | 12.84 | 0.10 |
| High blood pressure |  |  |  |  |
| Yes | 0.60 | 0.00 | 116.15 | 0.85 |
| No | 5.71 | 1.62 | 20.12 | 0.01 |
| PIR |  |  |  |  |
| <2.5 | 4.87 | 0.83 | 28.55 | 0.08 |
| >=2.5 | 1.08 | 0.04 | 27.04 | 0.96 |

Above adjusts for Age, Sex, Race, Education, Marital, PIR, Alcohol, High blood pressure, Diabetes, Creatinine level, Smoke.
In each case, the model was not adjusted for the stratification variable itself.

The nonlinear association between BLLs and bladder cancer was then described using smoothed curve fitting (Figures 2 and 3). Figure 2 shows a nonlinear relationship between BLLs and bladder cancer in the general population. Adjusted variables: age, sex, race, education, marital, PIR, BMI, alcohol, high blood pressure, diabetes, creatinine level, smoke. In people with a BMI<28kg/m^2^, a clear rising curve was detected, as shown in Figure 3. Adjusted variables: age, sex, race, education, marital, PIR, alcohol, high blood pressure, diabetes, creatinine level, smoke.

**Figure 2.**
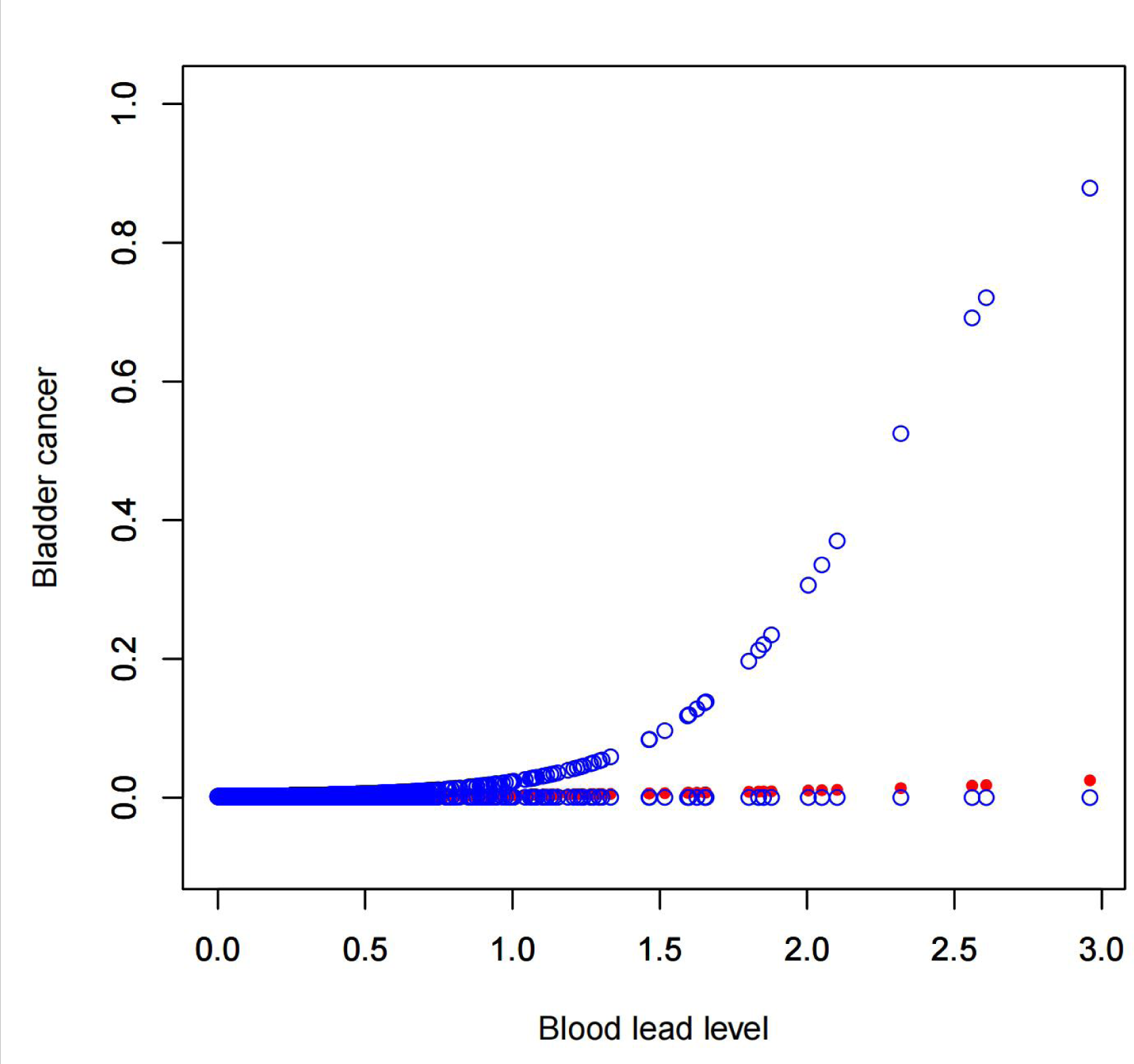
The association between BLL and bladder cancer, The solid red line represents the smoothcurve fit between variables. Blue bands represent the 95% confidence interval from the fit. Adjusted variables: age, sex, race, education, marital, PIR, BMI, alcohol, high blood pressure, diabetes, creatinine level, smoke.

**Figure 3.**
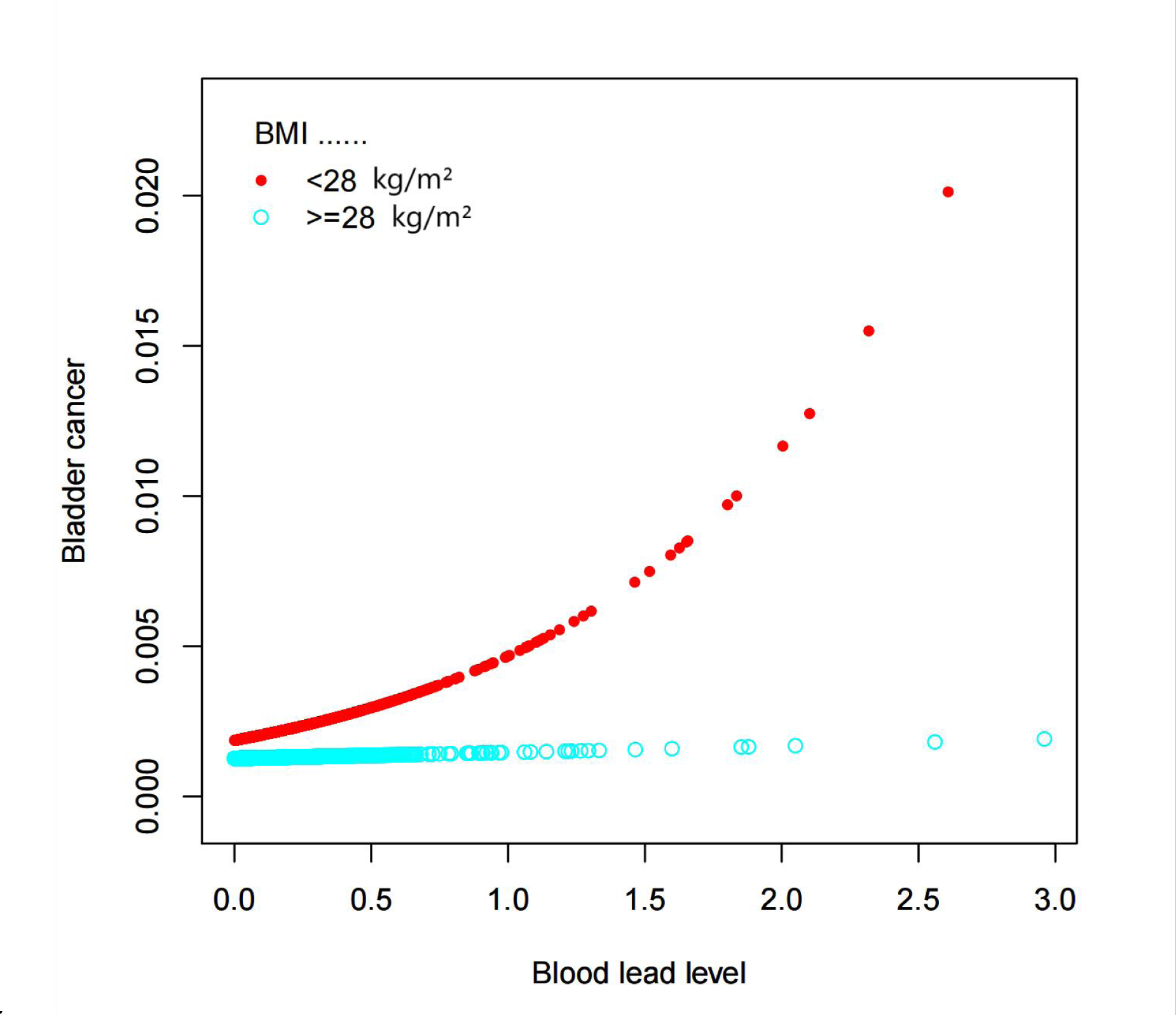
The association between BLL and bladder cancer stratified by BMI. The red line represents the smoothcurve of participants with BMI < 28kg/m^2^. Blue line represents the smoothcurve of participants with BMI≥28kg/m^2^. Adjusted variables:Age, Sex, Race, Education, Marital, PIR, Alcohol, High blood pressure, Diabetes, Creatinine level, Smoke.

## Discussion

In this cross-sectional study from NHANES 1999-2018, we identified higher BLLs was independently associated with Bladder cancer in people with BMI < 28kg/m^2^, demonstrated that blood lead is the risk factor for bladder cancer in this group, although there was no significant difference in general population. BMI> 28kg/m^2^ is defined as obesity, however we did not find a clear association between BLL and bladder cancer in obese people.

Lead is a heavy metal which can be found in many places. It can enter and accumulate in the human body through different ways. Accumulation in various organs will cause adverse reactions and may damage hematopoietic function, nervous system, cardiovascular system, reproductive system, urinary system, digestive system, etc(Ni et al. 2022; Qayyum et al. 2012; Rehman et al. 2018; Reja et al. 2020; Rhee et al. 2021; Tang et al. 2022; Zhu et al. 2022). It can lead not only to neuropsychiatric diseases but also to the progression of cancer(Ebrahimi et al. 2020). According to the International Agency for Research on Cancer (IACR), lead and its compounds are classified as “probable” human carcinogens (Group 2A)([Anonymous] 2006), and there are many articles revealing that lead may cause cancer. By measuring the chemical composition of breast cancer tissues and evaluating the DNA methylation level, a study involving 40 breast cancer tissues and 20 normal breast tissue found that the increase of heavy metals in tumor tissues was accompanied by the increase of the expression of HER2/neu, p53, Ki-67 and MGMT, and the decrease of the expression of ER and PR. Pathological DNA methylation increased with the increase of heavy metal content in tumor tissue, and the results showed that the average lead concentration in breast cancer tissue was almost three times that of healthy breast tissue, which means that heavy metals stimulate breast cancer progression and reduce its sensitivity to treatment(Romaniuk et al. 2017). A review suggests that lead concentrations may be associated with the development and progression of gynecological tumors such as cervical cancer, endometrial cancer, and ovaries(Furtak et al. 2022). In addition, Basu S et al. measured the levels of heavy metals in serum, bile, gallstones and gallbladder of 30 patients with gallbladder cancer and 30 patients with cholelithiasis by atomic absorption spectrophotometry and compared the measured results. It was found that the levels of selenium and zinc in serum, bile and gallbladder of patients with gallbladder cancer were significantly reduced, and the concentration of copper was significantly increased. Patients with gallbladder cancer have elevated levels of lead, cadmium, chromium and nickel in serum and bile(Basu et al. 2013). Taking 56 hair samples (aged 37 to 74 years) and 43 nail samples (aged 38 to 72 years) from untreated lung cancer patients, 54 scalp hair samples (aged 20 to 63 years) and 45 nail samples (aged 34 to 63 years) from healthy subjects. Then the heavy metal content of the samples was determined by flame atomic absorption spectrophotometer (Shimadzu AA-670, Japan), and the conclusion was finally drawn by Qayyum MA et al.: The majority of metals (Cd, Pb, Co, Ni, and Cu) in the scalp hair and nails of lung cancer patients were significantly higher than in the control group, and there was considerable variation in metal levels for different stages and types of lung cancer(Qayyum and Shah 2014a). In addition to pancreatic cancer, Qayyum MA et al. also clarified through the same method that the content of Pb in the blood and hair of prostate cancer patients was significantly higher than that of the control group(Qayyum and Shah 2014b). A systematic review and meta-analysis suggested that lead exposure was not associated with the risk of both benign and malignant brain tumors (pooled OR = 1.11, 95% CI: 0.95,1.29), but was associated with malignant brain tumors (pooled OR = 1.13, 95% CI: 1.04,1.24)(Ahn et al. 2020). Besides, in a study involving 188 exocrine pancreatic carcinoma and 399 control cases, Amaral AF et al. used inductively coupled plasma-mass spectrometry to determine trace element spectrometry for toenails. The results showed that the content of cadmium and lead in toenail was significantly higher than that in control group (P<0.001), and lead was associated with an increased risk of pancreatic excrine carcinoma (OR=6.26, 95%CI 2.71,14.47;Ptrend=3×10^−5^)(Amaral et al. 2012).

But there are also articles suggesting that lead is not clearly associated with some cancers. A case-control study investigated the occupational elements associated with lung cancer in 1593 male lung cancer patients and 1426 general population.Lifetime occupational exposure was assessed using expert-based blinded assessment, and adjustments were made for several potential confounding factors. Ultimately, it was concluded that exposure to lead compounds does not increase the risk of lung cancer(Wynant et al. 2013). Gaudet MM et al. conducted a meta-analysis with a large sample size and analyzed three case-control studies, and concluded that cadmium and lead levels in adults were not associated with an increased risk of breast cancer(Gaudet et al. 2019). For 1,217 cases of kidney cancer and 1,235 normal controls, experts used occupational history information to estimate occupational lead exposure. Unconditional logistic regression was used to calculate the odds ratios (ORs) and 95% confidence intervals (CIs) of different exposure indicators. This case-control study found that cumulative occupational lead exposure was not associated with kidney cancer (OR=0.9,95%CI 0.7,1.3; Ptrend=0.80)(Callahan et al. 2019). A cross-sectional study that also used the NHANES database analyzed 1,162 samples and used mediation analysis to examine the mediating effect of blood lead in the occurrence of cardiovascular diseases (CVD), respiratory diseases, and cancer. The study results did not find a statistically significant mediating effect(Yan et al. 2024).

In summary, whether lead causes cancer is still controversial, and there is very limited research on blood lead and bladder cancer, only one experimental study revealed a relationship between lead exposure and the initiation and development of bladder cancer. This study investigated the lead concentration levels in the blood and bladder cancer tissues of 36 bladder cancer patients, and compared them with 15 normal control groups. Atomic absorption spectroscopy was used to determine the lead concentration levels in each tissue sample. The results suggested that both the lead concentration levels in the cancer tissues and blood of bladder cancer patients were higher than those in the control group, while there was no relationship between the lead concentration levels in the bladder cancer tissues and the corresponding blood lead concentration(Golabek et al. 2009). Obviously, the sample size of this article is too small to be persuasive. Our paper fills the gap in the research field of lead and bladder cancer, and at the same time, the large sample size makes up for the shortcomings of previous studies, revealing higher blood lead level was associated with bladder cancer in people with BMI<28kg/m^2^.

It is unclear what biologic effects might underlie a causal association between lead exposure and bladder cancer, should one exist, here are the possible mechanisms. By replacing other ions such as Ca^2+^, Mg^2+^, Fe^2+^ and Na^+^, lead interferes with cellular metabolism(Flora et al. 2008), which indirectly leads to an imbalance in antioxidant responses(Kasperczyk et al. 2012) that affects key enzymes and hormones(Dhir et al. 2011). In addition to direct damage, lead increases the levels of reactive oxygen species and calcium ions in cells, reduces mitochondrial potential and reduces apoptosis through the release of cytochrome(Moreira et al. 2001). Continuous lead exposure may also affect immune function, which can induce cancer(Metryka et al. 2018). In addition, there are three more carcinogenic mechanisms: one is the activation of REDOX sensitive transcription factors, another involves their role as a mitotic signal, and the last one is lead interferes with the process of DNA repair(Genestra 2007; Silbergeld 2003).

Our study found that blood lead concentration was associated with bladder cancer in the general population, but the association became insignificant after accounting for covariates. Interestingly, blood lead was still found to be associated with bladder cancer in people with a BMI<28kg/m^2^, possibly because obesity represents a complex interaction of genetics, diet, metabolism, and physical activity levels, obese people have more risk factors than non-obese ones, the impact of lead on bladder cancer might be compensated by other confounding factors. In addition, a cross-sectional study(Park and Lee 2013) using the Korean National Health and Nutrition Examination Survey (KNHANES) database performed a multivariate linear regression analysis on 4,522 participants. The authors found a negative correlation between blood lead levels and body fat percentage and fat intake(Park and Lee 2012), suggesting that body fat percentage can be used as a predictor of blood lead, which partly explains our results. This interesting finding may suggest that lead may accumulate more in adipose tissue while the blood level is lower, resulting in underestimation of the systemic accumulation of lead. It is also possible that fat can improve the body’s tolerance to lead and may be a potential protective factor for lead carcinogenesis. Moreover, it can be seen from Table 2 that blood lead is negatively correlated with BMI, which further verifies our hypothesis. The specific reasons require further research and analysis.

Our research has the following advantages. Firstly, our study’s reliability and representativeness were enhanced for the data in this study were obtained from NHANES with a large sample size and covariates were considered to reduce confounding factors. Furthermore, our findings were validated by Weighted multivariate logistic regression and subgroup analysis to illustrate the risk of higher BLL on bladder cancer formation.

However, this investigation also has limitations. This was a cross-sectional study and the data came from an observational survey. It cannot prove causation, only association. Although we have tried to minimize confounding factors, there are still some limitations such as the use of single blood lead measurements, which may not accurately reflect cumulative lead exposure. In addition, the exact mechanisms behind this positive relationship are not well elucidated, because the causes of bladder cancer are extremely complex and are influenced by many genetic and environmental factors and there were still some unobserved confounding factors or information was not collected in the survey, more prospective studies should be done in the future.

## Conclusion

In conclusion, we discovered suggestive evidence of a relationship between increased BLLs and bladder cancer. Although the significant difference in general population disappeared after being adjusted for confounding factors, our findings demonstrated that bladder cancer risk increases with increased blood lead levels in people with BMI<28kg/m^2^. Because the findings were insufficient to show a causal association, more comprehensive prospective investigations are required.

## Data availability statement

Publicly available datasets were analyzed in this study. This data can be found here: https://www.cdc.gov/nchs/NHANES/index.htm.

## Ethics statement

The studies involving human participants were reviewed and approved by NCHS Research Ethics Review Board (ERB). Written informed consent for participation was not required for this study in accordance with the national legislation and the institutional requirements.

## Author contributions

Hongxiao Li conducted statistical analyses and Mei Huang wrote the manuscript. All the author worked together on the design of the study, supervised the project, conducted some statistical analyses, and critically reviewed the manuscript.

## Data Availability

https://www.cdc.gov/nchs/NHANES/index.htm

## Acknowledgments

The authors would like to express my sincere gratitude to the staff and participants of the NHANES study. Without their dedication and contribution, this research would not have been possible.

## Funding

Guangzhou Science and Technology Plan Project (Project No. 202201010834); Natural Science Foundation of Guangdong Province (Project No. 2021A1515010065); Guangzhou Health Science and Technology Project (Project No. 20211A011103); The Ministry of Education’s Industry-University Cooperative Education Project in 2022 (220904082210823);

Guangzhou Medical University Student Innovation Ability Improvement Plan in 2022 (106);

Doctoral Program of Guangdong Nature Foundation (2017A030310148)

